# Health-Related Quality of Life Perception Among Older Persons with Non-Communicable Diseases in Primary Healthcare Facilities: A Qualitative Inquiry

**DOI:** 10.1101/2024.07.19.24310704

**Authors:** Atim Fiona, Ndagire Regina, Chloe Nampima, Frank Kiyinji, Catherine Lwanira, Rose Clarke Nanyonga, Faustino Orach-Meza

**Author notes:** **Corresponding author:** Atim Fiona^1,2^, **Contact:**, **Email:**.

## Abstract

**Background:** Unveiling the understanding of older persons with non-communicable diseases (NCDs) regarding health well-being is paramount and can translate to increased self-efficiency, independence, and enhanced well-being. However, little is known about older persons’ understanding of the concept of health-related quality of life (HRQoL) in Uganda. The study explored perceptions of older persons with NCDs on HRQoL in central Uganda.

**Methods:** This exploratory qualitative study design involved 23 participants recruited from selected Primary healthcare facilities in Central Uganda. Thematic analysis using an inductive approach generated themes that informed the study’s qualitative findings.

**Results:** The key themes that emerged from the study include holistic well-being, lifestyle modification, and financial stability. The key component of HRQoL that came out clearly from the study was the physical domain. There is a need to embrace a person-centered approach based on the perceptions of older persons on HRQoL, which has the potential to improve well-being and enhance a healthy aging journey.

## Introduction

Health-related quality of life (HRQoL) is a crucial point of observation in public health that gained momentum in the last decade. HRQoL is a relevant predictor of prognostic significance (Haraldstad et al., 2019) employed to measure and identify health problems associated with dysfunctions related to physical and mental well-being. Literature shows that older persons live longer due to the increased life expectancy. However, they face enormous chronic conditions (WHO, 2017). Gaining an understanding of how older persons perceive HRQoL is key in guiding evidence-based mechanisms to enhance good physical and mental well-being. Much as ascertaining the perceptions of older persons on HRQoL is paramount in laying strategies for developing health policy frameworks (Jalenques *et al*., 2020; WHO, 2017). There are few studies done among older persons to unveil their perceptions towards HRQoL, with no single study done among older persons with non-communicable diseases (NCDs). Additionally, studies have centered on self-reported HRQoL without paying attention to the older person’s understanding of HRQoL concept (Jalenques *et al*., 2020; Dresden *et al*., 2019). The current study aimed to explore HRQoL perceptions among older persons with non-communicable to suggest strategies to enhance health well-being.

The World Health Organization defined HRQOL as an individual’s perception of their position in life in the context of the culture, and value systems in which they live and to their goals, expectations, standards, and concerns (WHO, 1997). Hernanadez-Segura et al., (2016) affirm that HRQOL has been conceptualized as a global tool that is used to assess the effects of a health condition of interest (Hernández-Segura et al., 2022) and has been used to measure the well-being of people, most especially elderly persons with Chronic conditions (Roemer, Emily J., West, Kesley L., Northrup, Jessica B., Iverson, Jana & Cho, 2016). HRQOL has been perceived as an individual’s or group’s perceived physical and mental health over time (CDC, 2021), and is perceived as a good measure of assessing the efficacy of strategies such as health interventions, health care, and public policy (JL et al., 2022). HRQOL was developed on functionalism and reflects the vital aspect of assessing and treating and not merely focusing on the biological, but also the psychological and social factors influencing the health status of individuals (Vette, 2010).

The concept of HRQOL encompassing the physical and mental domains is an important indicator of improved access to medical care and attention among the elderly. Self-perceptions about aging and HRQOL are key issues that need to be explored by healthcare workers during clinical sessions with the elderly. Studies conducted in developed countries (United States, Switzerland, China, Slovenia, and Turkey) indicate that HRQOL is a global phenomenon that should be explored more to facilitate the inauguration of mechanisms to improve older persons’ well-being (Dresden et al., 2019; Hou et al., 2020). Based on the literature reviewed, it is clear that a gap exists in ascertaining HRQoL among older persons with NCDs.

Non-communicable diseases pose a global challenge and are responsible for 63% of deaths globally (WHO, 2023). Annually, NCDs are responsible for 41 million deaths (WHO, 2023). Despite its strategic component to alleviate NCDs, morbidity and mortality data indicate the rising impact of NCDs in low-resource countries where 80% of deaths are due to cardiovascular diseases (Global status report, 2014). NCDs have long-term effects due to their duration, and these are further precipitated by environmental, physical, behavioral, and genetic factors (WHO, 2023), it is therefore paramount to assess the perceptions of older persons with NCDs on HRQoL to design strategies that can help to curb the negative impacts of NCDs on HRQoL.

In a low-income country like Uganda, the situation is particularly challenging. Overall, the prevalence of poor HRQoL in Central Uganda Luwero and Nakaseke Districts was estimated at 52% (Yaya et al., 2020), a figure close to the national prevalence of 59.6% (Maniragaba et al., 2018). Poor HRQoL exacerbates unhealthy days, limits activities, and elevates health symptoms among older persons with NCDs in these districts. This high prevalence may be linked to the substantial burden of NCDs among older persons in Central Uganda, reported at 28.5% (Siddharthan et al., 2021); Guwatudde et al., 2015). The evidence of the high burden of NCDs among older persons in Uganda with no studies done to examine the HRQoL perceptions among older persons with NCDs on HRQoL, prompted the current study to be conducted.

### Methodology

This was an exploratory qualitative study design conducted between January and February 2023. The study was conducted in Luwero and Nakaseke districts in central Uganda. The districts were purposively selected based on the high prevalence of NCDs registered in Nakaseke District in 2015, which was projected at 28.5% (Guwatudde et al., 2015; Chang et al., 2019; Siddharthan et al., 2021), a prevalence higher than the national prevalence of NCD of 27%. The study employed a focus group discussion (FGD) method of data collection, a total of three FGDs with 8 older persons in each group (both females & males) were conducted. The FGDs lasted between 45 to 60 minutes and were audio-recorded. Participants were reimbursed for their time with 10,000 shillings and refreshments were provided after the sessions.

For each FGD group, a trained research assistant served as a moderator and another researcher. A written FGD developed and pilot-tested by the researchers was used. The phrasing was modified after pilot testing without changing the original meaning of the content. Participants for the FGD were older persons aged 65 years and above screened and confirmed NCD patients attending PHC facilities in selected facilities in Nakaseke (Semuto Health Center IV) and Luwero (Kalagala Health Center IV) districts. Participants were selected using convenience sampling techniques. FGDs were conducted following an interview guide. All participants provided written consent before data collection, confidentiality and privacy were ensured.

Participants were informed about the objective of the study which was to explore perceptions of HRQoL. Recruitment of additional FGD groups stopped once transcripts were reviewed and saturation of themes was obtained. Ethical review and approval were sought from the Clarke International University Research Ethics Committee (CIUREC) under protocol no: CLARKE-2022-404 and clearance to conduct the study was obtained from the Uganda National Council for Science and Technology (UNCST) under study reference no: SS1528ES. Participants were informed about the dissemination of the study findings through publication which they consented to.

Data collected were entered into the Nvivo software version 20.2 to generate codes. Focus groups were coded and transcribed verbatim. Thematic analysis using an inductive approach generated themes that informed the study’s qualitative findings.

## Results

### Demographic characteristics of the respondents

This section presents the results of the qualitative study done among 23 study participants who constituted focus group discussions (FGDs). Of the 23 participants, 13 (56.7%) were aged between 65-74 years, 15 (65%) were females, and 10(43.5%) were Catholics as indicated in Table 1 (see appendix).

**Table 1:**
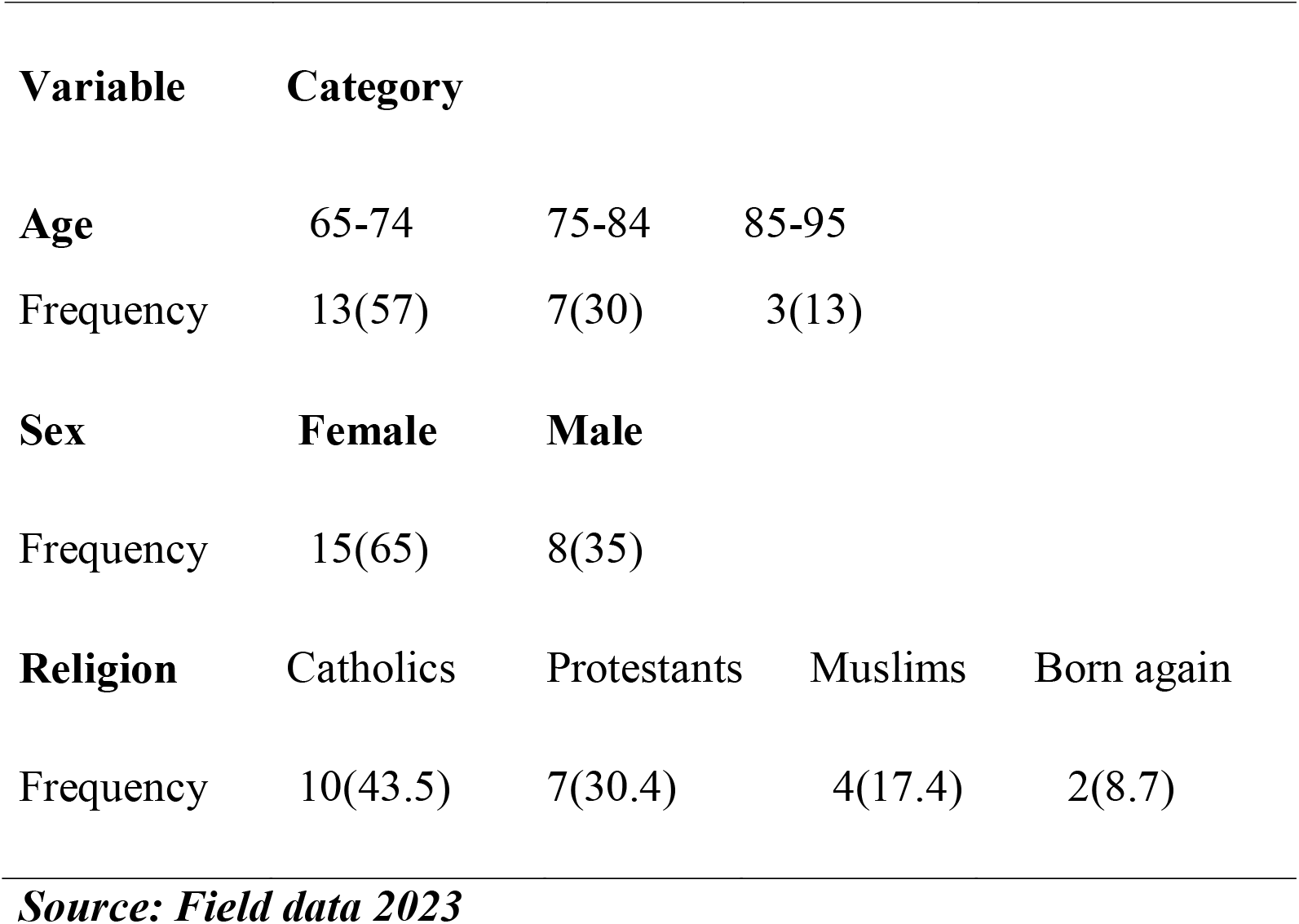
Demographic characteristics of the study respondents for FGD.

| Variable | Category |  |  |  |
| --- | --- | --- | --- | --- |
| Age | 65-74 | 75-84 | 85-95 |  |
| Frequency | 13(57) | 7(30) | 3(13) |  |
| Sex | Female | Male |  |  |
| Frequency | 15(65) | 8(35) |  |  |
| Religion | Catholics | Protestants | Muslims | Born again |
| Frequency | 10(43.5) | 7(30.4) | 4(17.4) | 2(8.7) |

***Source: Field data 2023***

### Subjective understanding of older persons on health-related quality of life

Through consensus, 14 codes were generated, and 3 codes related to HRQoL were categorized into themes. The 3 themes that emerged as older persons subjectively relayed their understanding of HRQoL: (1) holistic well-being, (2) lifestyle modification, (3) and financial stability. Themes with the corresponding verbatims in quotes were generated (see Table 2 in the appendix section) as described below with narrative examples.

**Table 2:** Additional Representative quotes.

| **Theme** | **Representation** |
| --- | --- |
| Holistic well-being | *“My ability to interact freely with my family and community, being healthy, having no disease, ability to support myself physically defines my HRQoL” Vb-6 source. FGD3/k/2023* |
|  | *“Having energy, absence of pain, participation in household chaos and garden work” Vb-7 source. FGD2/k/2023* |
|  | *“To me, HRQoL means having good health, no part of the body hurting, eating well, having a good quality of sleep”* |
| Lifestyle Modification | *“When I do everything required of me to keep healthy on time like taking my medicine, eating food, and staying away from things that will prevent the medication I take to work” Vb-14 source FGD1/S/2023* |
|  | *“I think it implies changing from unhealthy ways of life to more healthy ones” Vb-9 source FGD3/S/2023* |
| Financial stability | *“Having the money to meet my daily needs” Vb-20 source FGD2/S/2023* |
|  | *“On my side, having what it takes to maintain a good life, like having a daily source of income defines my HRQoL” Vb-17source FGD1/S/2023* |

**Source: Field data 2023**

### Theme 1: Holistic Well-being

Holistic well-being involves the complete state of health, including physical, mental, and social well-being. Participants noted that being physically healthy and able to perform daily duties, maintaining good mental health described and informed by the absence of worries and stress, and being supported and loved by family members: the majority of the participants pointed out support from a spouse and biological children, and having access to social health as the important aspects of HRQoL. The participants viewed HRQoL as having basic health, access to mental and physical, and social amenities. To some participants having access to medical care was reassuring.

> *“Health-related quality of life to me means being happy, no pain, no stress, good food, and support from family” (****Vb-8 source*. FGD1/S/2023)**

> *“Being healthy, having access to medical treatment, absence of disease, pain, care from my family, and having a community that relates well with people”* ***Vb-2 source*. FGD14/k/2023**

> *“Having what it takes to improve my life, Medicine, house, food, and having enough money to earn a living” Vb****-10 source*. FGD20/S/2023**

### Theme 2: Lifestyle Modification

The participants highlighted the importance of healthy living through behavioral change like regulating one’s diet, refraining from drug abuse, and engaging in physical activities like exercising and weight management. A common thought was as easy as “adjustments of harmful habits”

> *“I perceive health-related quality of life to be things that l do that make me healthy like avoiding alcohol intake and eating foods low in fats”* ***Vb-4 source*. FGD6/k/2023**

### Theme 3: Financial Stability

Participants discussed the importance of finances in enhancing good HRQoL. The majority of the participants asserted that money helps to facilitate transport, and buying medication in the event there is a drug stockout, a common phenomenon in public health facilities in Uganda. Financial stability was centered on good financial status and enhanced access to healthcare. Participants perceived financial status to have a positive impact on both individual and local economic development through the operation of small businesses and farming and went ahead to state that having access to finances from relatives, friends, and financial institutions through development loans and government funds for senior citizens improves HRQoL,

> *“Receiving constant assistance in the form of money from family, relatives, and friends makes me worry less. Having money promotes happiness, when l have money, I feel very healthy and active* ***Vb-17 source. FGDI2/k*/2023**

> *“Having access to government funds and pension as a retired senior citizen would define my health-related quality of life because l can use the money to start a business and be able to acquire assets like land”* ***Vb-20 source FGD7/S*/2023**.

## Discussion

The study used a qualitative approach to understand older people’s perceptions of HRQoL. The study found understanding of HRQoL to be characterized by holistic well-being, lifestyle modification, and financial stability. The key component of HRQoL that came out clearly from the study was the physical domain, which implies that older persons perceive physical well-being as an important issue in promoting well-being. Findings from previous studies are in line with the current findings where the majority highlighted the physical component of HRQoL to influence well-being compared to the mental component (Wang *et al*., 2023; Defar *et al*., 2023; Dresden et al., 2019; Lee, Cha and Kim, 2021). Other studies found common themes that were linked to older person perceptions of HRQOL in the previous study to include anxiety driven by emotional experiences, a strong desire to regain physical function, interpersonal impacts of health on quality of life driven by social aspects, socio-economic status denoted by high income, higher level of education, and individual experiences of physical and mental health status (Dresden *et al*., 2019; Lee, Cha and Kim, 2021). Furthermore, self-efficacy, numeracy in health literacy, physical health-promoting behavior, perceived support, and a lesser frequency of comorbidities were themes from a study conducted by Lee et al., (2021).

Findings from another study are divergent from the current findings, with poorer self-perceptions of HRQOL among elderly persons in rural areas being recorded compared to their counterparts in urban areas (Liu et al., 2020). A similar study conducted in Iran found self-reported HRQOL among older persons to be poor and characterized by poor mental health status (Tourani et al., 2018). A discrepancy in findings from middle-income and low-income countries indicates that HRQOL was perceived to be driven by poverty, lack of social support, psychological status, and negative self-perceptions of aging (Maniragaba *et al*., 2018; Yaya *et al*., 2020; Craig *et al*., 2016). A study done in Uganda by JL et al., (2022) shows that older persons had positive perceptions towards HRQOL which is similar to the current findings whereby only positive HRQoL was recorded. However, much as the previous study reported positive HRQoL, it did not stop the participants from having poor HRQOL as 86% of the respondents had poor HRQOL when assessed quantitatively.

Some of the issues noted to influence the perceptions of older persons towards their HRQOL include lack of social and financial support, poor housing, and loneliness (JL, 2022; Dresden *et al*., 2019). The differences in findings between developed countries and underdeveloped countries may be attributed to the better socio-economic status and development of the aging population in developed countries compared to low-income countries. Another observation elucidates that older people in developed countries have high mental and psychological issues which can be attributed to loneliness since most of the older persons live by themselves which psychologically impacts their mental health.

Plotnikoff et al., (2015) argue that regular physical activity, and improved health interventions like maintaining a healthy body weight and refraining from a sedentary lifestyle influence health decisively and promote HRQoL (Plotnikoff et al., 2015). Addressing both the physical and mental health aspects of HRQoL begins with the healthcare workers’ knowledge and inquiry (Syamlan et al., 2022). Understanding older persons’ perceptions of HRQoL helps to design health interventions targeting improvements in HRQoL. At the same time, it shows that older persons know what HRQoL implies, which was one of the main focus of the current study. In addition to informing primary healthcare-based geriatric patient care, the study’s findings may be most applicable in HRQoL status evaluations in the future.

The current study reported a positive perception of HRQoL among older persons, but it had some limitations. First, participants were recruited from PHC facilities in two selected districts of central Uganda. Therefore, the limited sample size and sampling methods limit generalizability to a larger population in central Uganda. Secondly, the focus group guide was well-articulated hindering leading questions. However, HRQoL being a new term to older participants, some responses may have been steered by the carefully designed question guide. Thirdly, since the participants gave their subjective opinions concerning HRQoL, it is possible that some participants gave socially desirable responses to the moderators to please them. Despite these limitations, FGDs are an important qualitative research approach to evaluate concepts, such as how older persons conceptualize HRQoL.

## Data Availability

All data produced in the present study are available upon reasonable request to the authors

## Conclusions and Recommendations

The participants’ perceptions towards HRQoL were positively characterized by holistic well-being, lifestyle modification, and financial stability. Healthcare workers and policymakers should involve older persons in the planning and implementing interventions and policies that aim at creating awareness of HRQoL to enhance understanding of HRQoL concept to foster good health practices. Additionally, more studies need to be done to ascertain where subjective understanding of the study participants influences HRQoL outcome. There is a need to embrace a person-centered approach based on the perceptions of older persons on HRQoL, which has the potential to improve well-being and enhance a healthy aging journey. Given the limited qualitative research on aging, specifically perceptions of older persons with NCDs, more studies need to be done to enrich the scanty body of literature.

### Ethical considerations

This study was reviewed and approved by the Clarke International University Research Ethics Committee (CIUREC) approval number CLARKE-2022-404 and the National Council for Science and Technology (UNCST) clearance number SSI528ES. All participants consented to the study before participation.

## Members Contribution

All members actively contributed to the study. AF conceptualized the study and participated in writing the proposal, analysis, and report writing, NR participated in data analysis and manuscript writing; CN participated in data analysis; FR and CL guided the manuscript writing process and content; FOM and RCN supervised the entire research process as the study supervisors.

## Definition of key terms

CDC: Centre for Disease Control and Prevention
HRQoL: Health-related Quality of Life
NCDs: Non-communicable Diseases
PHC: Primary Healthcare

